# Secretory acid sphingomyelinase activity is elevated in persons with colorectal neoplasia

**DOI:** 10.64898/2026.01.22.26344557

**Authors:** Justin M. Snider, Elise K. Batzli, Madison L. Hannan, Aki Hara, Qiuming Wang, Juanita L. Merchant, Xavier Llor, Rosa M. Xicola, Elizabeth T. Jacobs, Peter Lance, Nathan A. Ellis, Ashley J. Snider

## Abstract

**Background:** Metabolomic changes related to colorectal cancer (CRC) may serve as diagnostic markers to identify patients may develop or have developed CRC.

**Methods:** Untargeted lipidomics were performed on serum from CRC cases and clean-colon controls from the Chicago Colorectal Cancer Consortium (CCCC) and the University of Arizona Cancer Center (UACC).

**Results:** Untargeted lipidomics in the CCCC CRC series revealed significant alterations in sphingolipids. Targeted lipidomics revealed a signature of five sphingomyelins (SMs) were significantly decreased in CRC patients in CCCC and UACC CRC series. Circulating SMs are degraded primarily by S-SMase and serum S-SMase activity was significantly higher in UACC cases as compared to controls. Serum S-SMase activity was also measured in two series of adenoma patients to determine if S-SMase may serve as a biomarker for development of colorectal neoplasia. While S-SMase activity was significantly higher in adenoma patients compared to controls in the mostly white UACC series, S-SMase activity in samples from the Chicago Black series (CCCC) were indistinguishable from each other and significantly higher than UACC controls.

**Conclusions:** Together, these studies suggest the potential for S-SMase activity to serve as a biomarker for colorectal neoplasia, with potential implications in some but perhaps not all populations.

## Introduction

Colorectal cancer (CRC) is the third most common cancer worldwide, with approximately 1.15 million new cases diagnosed and 576,00 new deaths per year [1]. In the United States, there will be an estimated 53,000 deaths in 2025 due to CRC, making it the second leading cause of cancer-related deaths. The incidence of CRC has been declining since the mid-1980s, most dramatically in adults over 65, widely attributed to increased CRC screening. However, since 1995, the incidence of CRC in persons under 50 years old, referred to as early-onset (EO)-CRC, has increased approximately 2% each year [2, 3], resulting in more than 18,000 EO-CRC diagnoses annually. In addition, incidence of and mortality from CRC is higher in black Americans than in non-Hispanic whites, and the incidence of EO-CRC has been increasing among non-Hispanic whites and in Hispanic patients [4] disproportionately relative to the overall incidence rate [5, 6].

In response to the rising incidence of EO-CRC, screening for CRC is now recommended at 45 years of age by the United States Preventative Services Task Force (USPSTF) [7, 8] and the American Cancer Society [9]. There are several practical issues inherent in this recommendation. First, based on the 2020 US Census, the recommendation adds over 14 million Americans to screening, and capacity in the healthcare system to screen these additional patients will need to be increased, potentially exacerbating the existing deficiencies in access to CRC screening. Secondly, the national screening rate overall in 2020 was 69.7%, but screening rates among persons aged 50-54 are below 50%, indicating that compliance with the recommendations for younger age groups is a serious challenge. Finally, the cost of screening, both in terms of out-of-pocket expenses and potentially unpaid time away from work, are burdens that affect lower-income patients more than higher-income patients. Thus, for all these reasons, there is an urgent need for blood- or urine-based risk assessment and screening tools to test for the presence of neoplasia in the colorectum. The recent FDA approval of a blood-based, cell-free DNA screening test [10] is evidence of this need, even though the reported sensitivity for CRC was 83.1% and for advanced precancerous lesions only 13.2%.

Discovery metabolomics, lipidomics, and other unbiased approaches are currently being explored to define molecular signatures to fill critical roles in disease diagnosis and prognosis. A recent study geared at examining lipidomics in tumor and adjacent normal tissues demonstrated a tumor-specific lipid signature based on glycerolipids, triacylglycerols, and sphingolipids [11]. A similar study examining plasma metabolites in CRC patients and healthy controls demonstrated alterations in phosphatidylglycerol, lyso-phosphatidylcholine, and phosphatidylethanolamine, as well as three sphingolipids [12]. These studies suggested that lipid metabolism is altered in both tumor tissues and circulation of patients with CRC; however, the potential for specific lipids to serve as part of a non-invasive risk assessment signature in CRC is yet unknown.

Sphingolipids are bioactive molecules that regulate numerous central events in the development and progression of cancer, including proliferation, apoptosis, angiogenesis, cell migration, and resistance to chemotherapeutics [13, 14]. Circulating sphingolipids are beginning to be appreciated as potential biomarkers in a variety of disease states, including CRC [15]. We conducted untargeted lipidomics on serum from CRC patients compared to persons with endoscopically confirmed absence of colorectal neoplasia and identified alterations in circulating lipid levels, specifically sphingolipids in CRC patients. Our analysis showed 13 species of sphingolipids, including of seven sphingomyelin (SM) species were decreased in CRC patients as compared to controls, and this decrease was strongly associated with increased levels of secretory acid sphingomyelinase (aSMase, S-SMase) activity in serum. Moreover, S-SMase activity from patients with adenomatous polyps in the colorectum increased in a predominantly white series of patients but not in a series of American Blacks. Taken together, this study suggests that increased S-SMase activity may be associated with the development of benign neoplasia in the colorectum and could serve as a biomarker for colorectal neoplasia in specific populations.

## Materials and Methods

### Patient Samples

To carry out an unbiased lipidomics analysis, we assembled a series of serum samples from CRC patients and controls. Serum samples were collected from five major medical centers in Chicago between 2011 and 2012 as part of the Chicago Colorectal Cancer Consortium (CCCC). Patients were identified in both endoscopy and surgical clinics as described by Xicola *et al*. [16]. Serum samples were obtained from patients with adenocarcinoma of the colon or rectum, with adenomatous colon polyps, or with no evidence of disease. Patients with polyposis or inflammatory bowel disease were excluded. Clinical, demographic, and epidemiologic data were collected and linked to the specimens. Serum samples were prepared as described and stored frozen at -80 °C. We assembled a CRC case-control series from these samples (CCCC CRC series, Table 1), consisting of CRC cases (n=61) and of controls with no colorectal neoplasia, endoscopically confirmed (referred to hereafter as clean-colon controls; n=47). To test promising leads, we assembled a validation series consisting of serum samples from 88 CRC cases from the University of Arizona Cancer Center (UACC) Tissue Acquisition Cell and Molecular Analysis Shared Resource (TACMASR). For comparison, we obtained serum samples from 17 clean-colon controls through the gastrointestinal clinic repository TARGHETS (Tissue Acquisition Repository for Gastrointestinal and Hepatic Systems; UACC CRC series, Table 2).

**Table 1.**
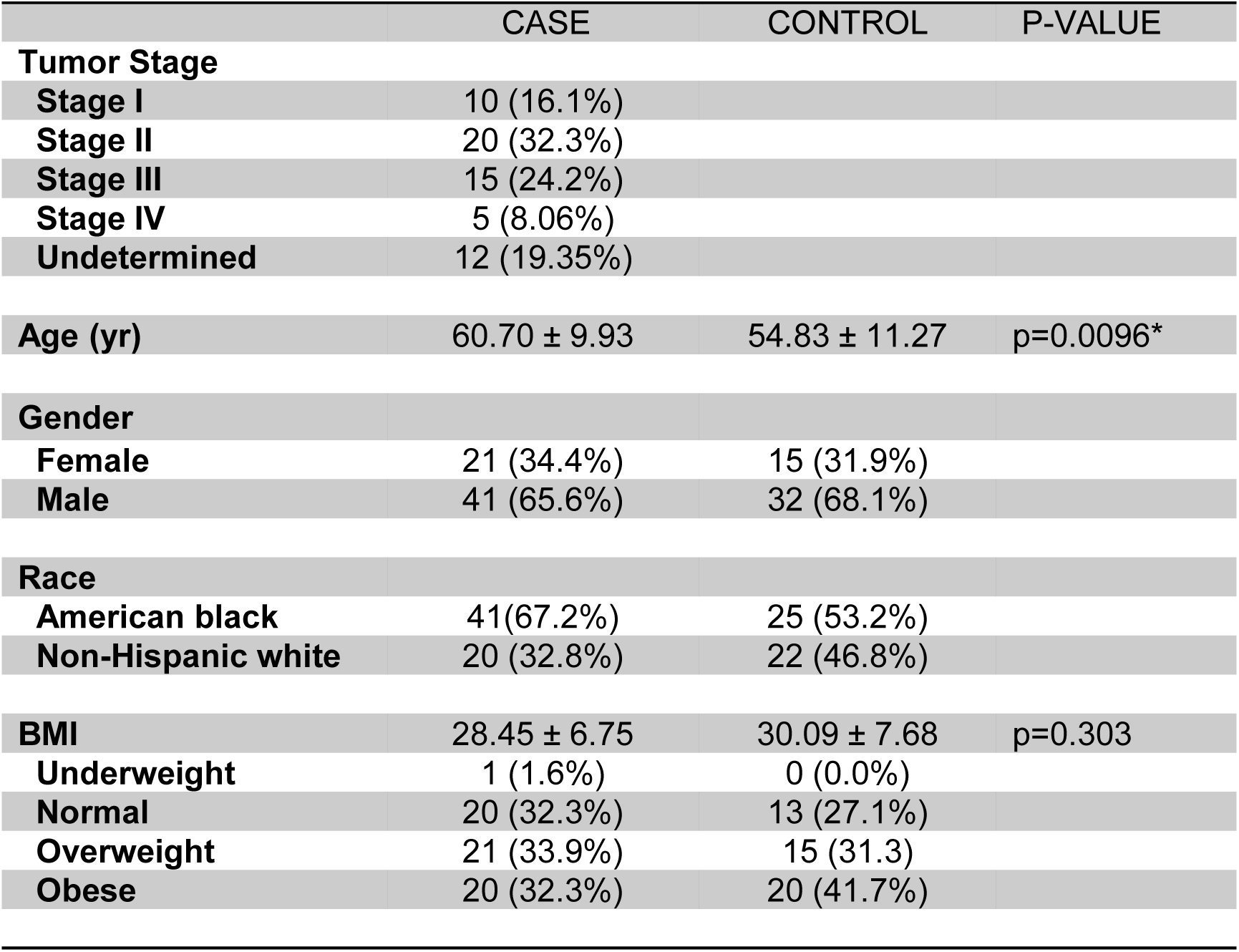
Demographics from the CCCC CRC Series.

|  | CASE | CONTROL | P-VALUE |
| --- | --- | --- | --- |
| **Tumor Stage** |  |  |  |
| **Stage I** | 10 (16.1%) |  |  |
| **Stage II** | 20 (32.3%) |  |  |
| **Stage III** | 15 (24.2%) |  |  |
| **Stage IV** | 5 (8.06%) |  |  |
| **Undetermined** | 12 (19.35%) |  |  |
| **Age (yr)** | 60.70 ± 9.93 | 54.83 ± 11.27 | p=0.0096* |
| **Gender** |  |  |  |
| **Female** | 21 (34.4%) | 15 (31.9%) |  |
| **Male** | 41 (65.6%) | 32 (68.1%) |  |
| **Race** |  |  |  |
| **American black** | 41(67.2%) | 25 (53.2%) |  |
| **Non-Hispanic white** | 20 (32.8%) | 22 (46.8%) |  |
| **BMI** | 28.45 ± 6.75 | 30.09 ± 7.68 | p=0.303 |
| **Underweight** | 1 (1.6%) | 0 (0.0%) |  |
| **Normal** | 20 (32.3%) | 13 (27.1%) |  |
| **Overweight** | 21 (33.9%) | 15 (31.3) |  |
| **Obese** | 20 (32.3%) | 20 (41.7%) |  |

**Table 2.** Demographics from the UACC CRC Series.

|  | CASE | CONTROL | P-VALUE |
| --- | --- | --- | --- |
| **Number** | 88 | 17 |  |
| **Tumor Stage** |  |  |  |
| **Stage I** | 21 |  |  |
| **Stage II** | 30 |  |  |
| **Stage III** | 30 |  |  |
| **Stage IV** | 7 |  |  |
| **Undetermined** | 0 |  |  |
| **Age (yr)** | 66.13 ± 12.71 | 50.76 ± 15.46 | 0.0002* |
| **Gender** |  |  |  |
| **Female** | 48 (54.5%) | 11 (64.7%) |  |
| **Male** | 40 (45.5%) | 6 (35.3%) |  |
| **Race** |  |  |  |
| **Non-Hispanic White** | 69 (78.4%) | 8 |  |
| **Hispanic** | 11 (12.5%) | 1 |  |
| **Native American** | 3 (3.4%) | 2 |  |
| **Asian** | 1 (1.1%) | 0 |  |
| **Mixed** | 0 (0.0%) | 5 |  |
| **Refused/Unknown** | 4 (4.5%) | 1 |  |

To determine whether S-SMase activity could be a biomarker for colorectal neoplasia, we assembled serum samples from two series of adenoma patients. The first series consisted of 126 patients ascertained for an adenoma recurrence trial. 77 had a non-advanced adenoma at baseline and a non-advanced or no adenoma at follow-up (low-risk) and 49 had an advanced adenoma at baseline and an advanced adenoma at follow-up (high-risk). Cases with advanced adenoma were defined as having an adenoma >= 1 cm or with villous features or with high-grade dysplasia, sessile serrated polyps >= 1 cm, a serrated lesion with any grade of cytologic dysplasia, or a traditional serrated adenoma regardless of size. 30 controls without a colorectal complaint were ascertained through the emergency room at Banner-University Medical Center (UACC adenoma series, **Table 3**; colonoscopy results not available for these controls). A second series of adenoma cases consisted of 30 screening patients with a non-advanced adenoma, 30 patients with an advanced adenoma, and 32 clean-colon control patients (CCCC adenoma series, **Table 4**). Race was self-reported in both CCCC and UACC patients. These studies were conducted according to the corresponding approved IRB protocols at each institution.

**Table 3.** Demographics from the UACC adenoma Series.

|  | HIGH RISK | LOW RISK | ALL CASES | CONTROLS | P-VALUE |
| --- | --- | --- | --- | --- | --- |
| **Number** | 49 | 77 | 126 | 30 |  |
| **Age (yr)** | 64.74 | 62.43** | 63.52 | 70.33 | p<0.01 |
| **Gender** |  |  |  |  |  |
| **Female** | 18 | 23 | 41 | 15 |  |
| **Male** | 29 | 30 | 59 | 15 |  |
| **Unknown** | 2 | 24 | 26 | 0 |  |
| **Race** |  |  |  |  |  |
| **Non-Hispanic White** | 47 | 48 | 95 | 24 |  |
| **Hispanic** | 0 | 3 | 3 | 6 |  |
| **Native American** | 0 | 0 | 0 | 0 |  |
| **Asian** | 0 | 1 | 1 | 0 |  |
| **Mixed** | 0 | 0 | 0 | 0 |  |
| **Refused/ Unknown** | 2 | 25 | 27 | 0 |  |

**Table 4.** Demographics from the CCCC adenoma Series.

|  | HIGH RISK | LOW RISK | ALL<br>CASES | CONTROL<br>S | P-VALUE |
| --- | --- | --- | --- | --- | --- |
| **Number** | 30 | 30 | 60 | 32 |  |
| **Age (yr)** | 64.9 | 62.5 | 63.7 | 63 | n.s. |
| **Gender** |  |  |  |  |  |
| **Female** | 15 | 15 | 30 | 15 |  |
| **Male** | 15 | 15 | 30 | 17 |  |
| **Race** |  |  |  |  |  |
| **American Black** | 30 | 30 | 60 | 32 |  |

### Sample Preparation

Fifty µL of serum were extracted with 190 µL of liquid chromatography mass spectroscopy (LCMS) grade methanol containing 10 µL of SPLASH LipidoMIX internal standard (IS) (Avanti, Al). Samples were vortexed (4°C for 30 minutes), centrifuged (13,000 RPM at 4°C for 15 minutes), and evaporated under nitrogen before final resuspension in 100 µL of LCMS grade methanol. Quality Control (QC) mixtures were created by pooling 10 µL from each sample.

### Un-targeted Lipidomic UPLC-MS Analysis

Serum samples were extracted and analyzed by Bioanalysis and Omics (ARC-BIO) at Colorado State University as previously described [17]. Briefly, serum extract was dried under nitrogen and resuspended in 100 μL of toluene/methanol (3/2, v/v). 3 μL of extract was injected onto a Waters Acquity Ultra-high performance liquid chromatography (UPLC) system in randomized order and separated using a Waters Acquity UPLC CSH Phenyl Hexyl column (1.7 µM, 1.0 x 100 mm), using a gradient from solvent A (water, 0.1% formic acid) to solvent B (Acetonitrile, 0.1% formic acid). Injections were made in 100% A, held at 100% A for 1 min, ramped to 98% B over 12 minutes, held at 98% B for 3 minutes, and re-equilibrated utilizing a 200 µL/min flow rate. The column and samples were held at 65°C and 6°C, respectively. Eluent was infused via electron spray ionization (ESI) source into a Waters Xevo G2 Quadrupole Time-of-Flight Mass Spectrometry (Q-TOF-MS) in positive mode, scanning 50-2000 m/z at 0.2 seconds per scan, and alternated between MS (6 V collision energy) and MSE mode (15-30 V ramp). Calibration was performed using sodium iodide with 1 ppm mass accuracy. Capillary voltage was 2200 V, source temperature was 150°C, and nitrogen desolvation temperature was 350°C with flow rate 800 L/hr. Annotations were assigned based on computational interpretation of MS signals.

### Targeted mass spectrometry analysis of sphingolipids

Serum sphingolipids were measured via liquid chromatography–tandem mass spectrometry (LC-MS/MS) as described previously [18, 19] with minor modifications. In brief, lipid extracts from each sample containing IS were dried under nitrogen, resuspended, and injected onto the TSQ Quantum Ultra LC-MS/MS system. Lipids were gradient eluted from the Spectra C8SR column (150 × 3.0 mm, 3-f particle size) identical to Bielawski et al. [18]. Peaks that corresponded to the target analytes and ISs were identified and processed using LCQuan software (Thermo Fisher Scientific, Waltham, MA, USA). Quantitative analyses of endogenous sphingolipids were based on calibration curves generated by ISs (Avanti Polar Lipids, Alabaster, AL, USA). For the analytes for which there was not a commercially available standard, the closest by mass and structure standard was used for quantification. Target analyte/IS peak area ratios were compared with the calibration curves by using a linear regression model. Levels of specific sphingolipids were normalized to volume of serum analyzed.

### S-SMase activity assay

S-SMase activity assay protocols were adapted from Jenkins *et al.* [20]. Briefly, 50 µl of 20 µM SM-d18:1/6:0 and 10 µl of 20 µM Cer-d18:1/4:0 (ISs) were dried under nitrogen and reconstituted in 100 µl of 250 mM sodium acetate (pH5.0), 0.2% Triton X-100, and 0.2 mM ZnCl_2_, utilizing sonication at 50 °C. 15 µl of serum was added to the reconstituted lipid mix and incubated at 37 °C for 4h at a shaker speed of 350 RPM. To quench reaction and extract the lipids, 1 ml of 2:3 70% isopropanol:ethylacetate was added to reaction mix. Lipids were then extracted and analyzed following adapted procedures from Beilawski et. al. [21]. Analysis was performed on a Thermo Quantum Ultra triple quadrupole mass spectrometer tandem Agilent 1200 LC System.

### Data Analysis and Statistics

For each untargeted lipidomics sample, raw data files were converted to cdf format, and a matrix of molecular features defined by retention time and mass (m/z) was generated using XCMS software in R [22] for feature detection and alignment. The centWave algorithm was used for LC-MS data. Features were grouped using RAMClustR [23], with normalization set to total ion current. LC-MS data were annotated by searching against an in-house spectra and retention time database using RAMSearch. RAMClustR was used to call the findMain [24] function from the interpretMSSpectrum package to infer the molecular weight of each LC-MS compound and annotate the mass signals. The complete MS spectrum and a truncated MSE spectrum were written to .mat format for import to MSFinder [25]. The MSE spectrum was truncated to only include masses with values less than the inferred M plus its isotopes, and the .mat file precursor ion was set to the M+H ion for the findMain inferred M value. These mat spectra were analyzed to determine the most probable molecular formula and structure. MSFinder was used to perform a spectral search against the MassBank database. All results were imported into R and a collective annotation was derived with prioritization of RAMSearch > MSFinder mssearch > MSFinder structure > MSFinder formula > findMain M. Annotation confidence was reported as described [26]. All R work was performed using R 3.3.1 [27]. Classical univariate receiver operating characteristic (ROC) curves, as well as all statistical analysis of untargeted data were generated using Metaboanalyst 6.0 [28]. For categorical data, statistical analyses were conducted using GraphPad Prism version (10.4.2). Grubb’s tests were utilized for outlier detection. In normally distributed data, an unpaired Student’s t-test or One-way ANOVA were performed, otherwise Mann-Whitney and Kruskal-Wallis tests were performed, with false discovery rate (FDR) correction. Multiple linear regression analyses were conducted using GraphPad Prism (10.4.2) using two models: 1) SM or S-SMase ∼ intercept + age + sex + race + affection status (case/control) or S-SMase ∼ intercept + age + sex + risk (low/high/control). A p-value < 0.05 was considered significant.

## Results

### Levels of serum sphingomyelins are decreased in CRC patients

To identify metabolites associated with CRC, untargeted lipidomics were conducted at the Colorado State University Bioanalysis and Omics facility using CCCC serum samples from newly diagnosed CRC cases (n=62) and clean-colon controls (n=48) (**Table 1**). To construct a list of potentially discriminating analytes, we utilized statistical software specifically designed for untargeted mass spectrometry data. First, univariate methods with t-test (p<0.05) and fold change (Log2 Fold Change > 1.3) identified eight molecules with significantly increased relative abundance in CRC patients and 32 molecules with significantly decreased relative abundance (**Figure S1A**). Secondly, multivariate, machine learning models, were conducted including linear and non-parametric methods, selected metabolites high variable importance in projection (VIP) scores by partial least squares-discriminant analysis (**Figure S1B-D**). Importantly, cross-validation indicated an accuracy of greater than 0.7, indicating the model was not over-fitted. Lastly, random forest was implemented to predict the importance of metabolites with the greatest mean decrease in accuracy (**Figure S1E**). Top hits from all three analyses identified 48 metabolites that were significantly different between case and controls. These metabolites spanned six major classifications of metabolites (**Table S1**). Thirteen of the 48 metabolites on the list were sphingolipids, including seven species of SM and two ceramide phosphoethanolamines (PE-Cer). Of the five glycerolipids, two triacylglycerols (TAGs) and one diacylglycerol (DAG) were identified (**Table S1**). These data demonstrated that specific serum lipids are significantly different between CRC patients and controls.

Due to the inherent semiquantitative nature of untargeted lipidomics, we utilized targeted liquid chromatography and tandem mass spectrometry (LC-MS/MS) to quantify and confirm sphingolipid levels, as these represented a substantial fraction of the candidate metabolites that were different between cases and controls. Absolute levels of five species of SM (C14:0, 20:0, 20:1, 22:0, and 22:1) were measured and all were significantly decreased in serum from CRC cases versus controls (**Figure 1A-E**) (Note: d18:1/18:2 and d18:1/23:0 were not analyzed due to very low abundance and lack of authentic standards). As expected, the signature SMs (sum of five identified significant species) were also significantly decreased in cases versus controls (**Figure 1F**). These data substantiated the untargeted lipidomics and further demonstrated significant decreases in specific SMs in serum from CRC patients.

**Figure 1.**
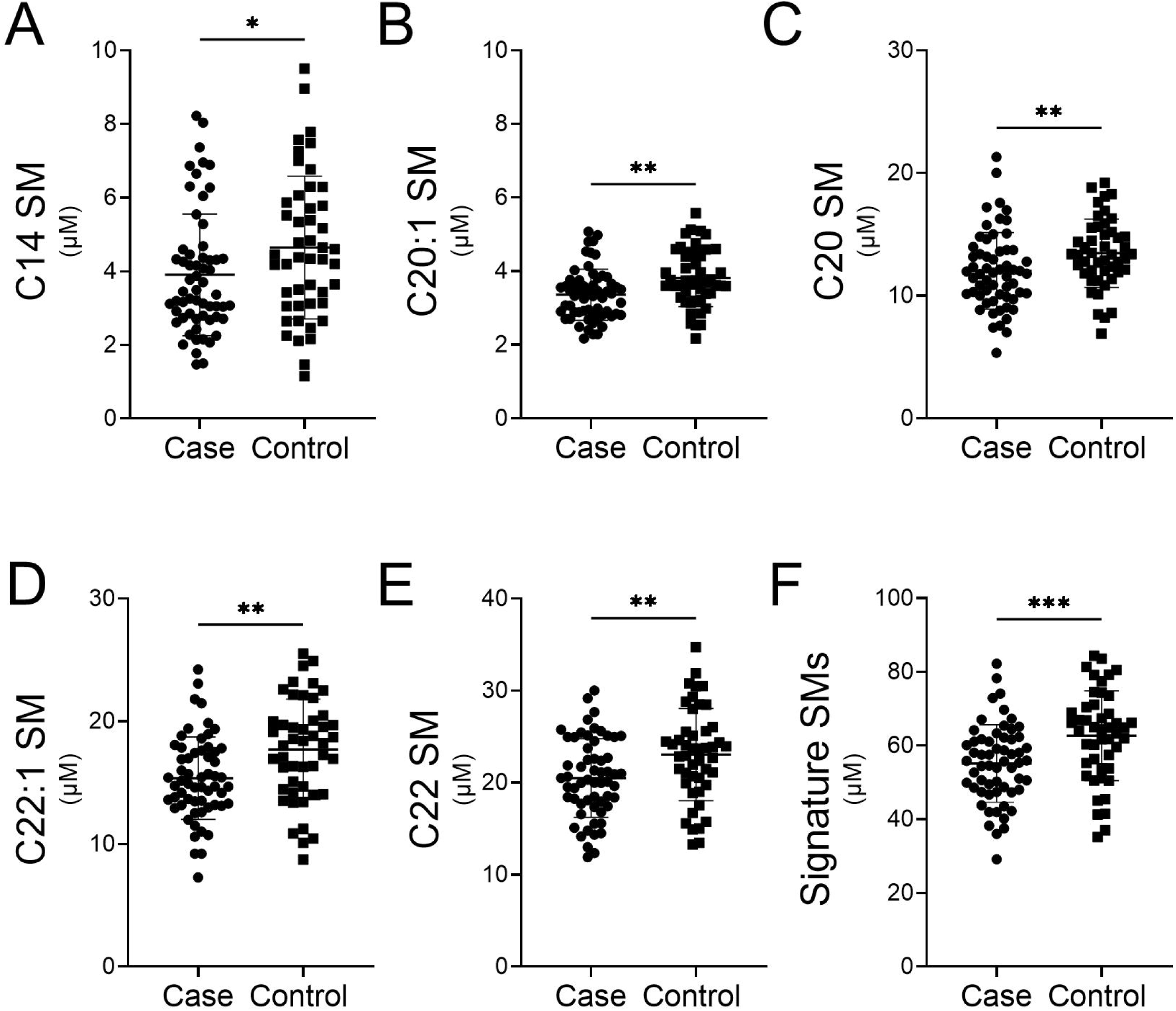
Signature SMs are decreased in CRC patients from CCCC series. Lipids in serum from CCCC CRC patients and clean-colon controls were extracted and analyzed for sphingolipids using LC-MS/MS. Five SM species identified in the untargeted lipidomics were quantified **A)** C14, **B)** C20:1, **C)** C20, **D)** C22:1, **E)** C22, **F)** sum of signature SMs (C14, C20:1, C20, C22:1). Data are normalized to volume and represent mean ± SD; *p<0.05, **p<0.01, ***p<0.0001.

Based on multiple linear regression analyses, sex was significant associated with levels of signature SMs (*P*=0.0076, 95% CI: [1.707, 10.86]; **Table S2**), whereas race was not; age trended in the model (*P*=0.052). Secondary stratification of categorical targeted MS data revealed significant differences in C14:0 SM among male cases versus controls, and C20:1 and C22:1 among female cases versus controls (**Figure S2A-F**). Signature SM levels were significantly decreased in both male and female cases in comparison to controls (**Figure S2G**). Two of the five SMs (C20:1 and C22:1), as well as signature SMs were significantly decreased in Chicago Black CRC patients as compared to Chicago Black controls (**Figure S3A-G**). No significant differences were measured in any of the SMs in non-Hispanic whites, including signature SMs (**Figure S3**).

To validate alterations in serum SMs in CRC cases compared to clean-colon controls, we again used targeted LC-MS/MS to measure SM levels in a second, independent series from the UACC (**Table 2**). Three of the original five SMs (C20, C22:1, and C22) that were decreased in CCCC CRC cases were also significantly lower in UACC CRC cases (**Figure 2A-E**). Similarly, signature SM levels were significantly lower in UACC CRC cases compared to clean-colon controls (**Figure 2F**). Additional targeted analyses determined that sphingosine- and sphinganine-1-phosphate were significantly elevated in cases vs. controls, and two additional SMs (C24 and C24:1) were decreased in cases vs. controls (**Table S3**). Based on multivariate analyses, sex again was significantly associated with the levels of signature SMs in the UACC CRC series (*P*=0.0004, 95% CI: [3.987, 13.21]; **Table S4**), whereas age and race were not. The validation of the targeted SM analyses demonstrated the potential for specific serum SMs to serve as biomarkers for CRC.

**Figure 2.**
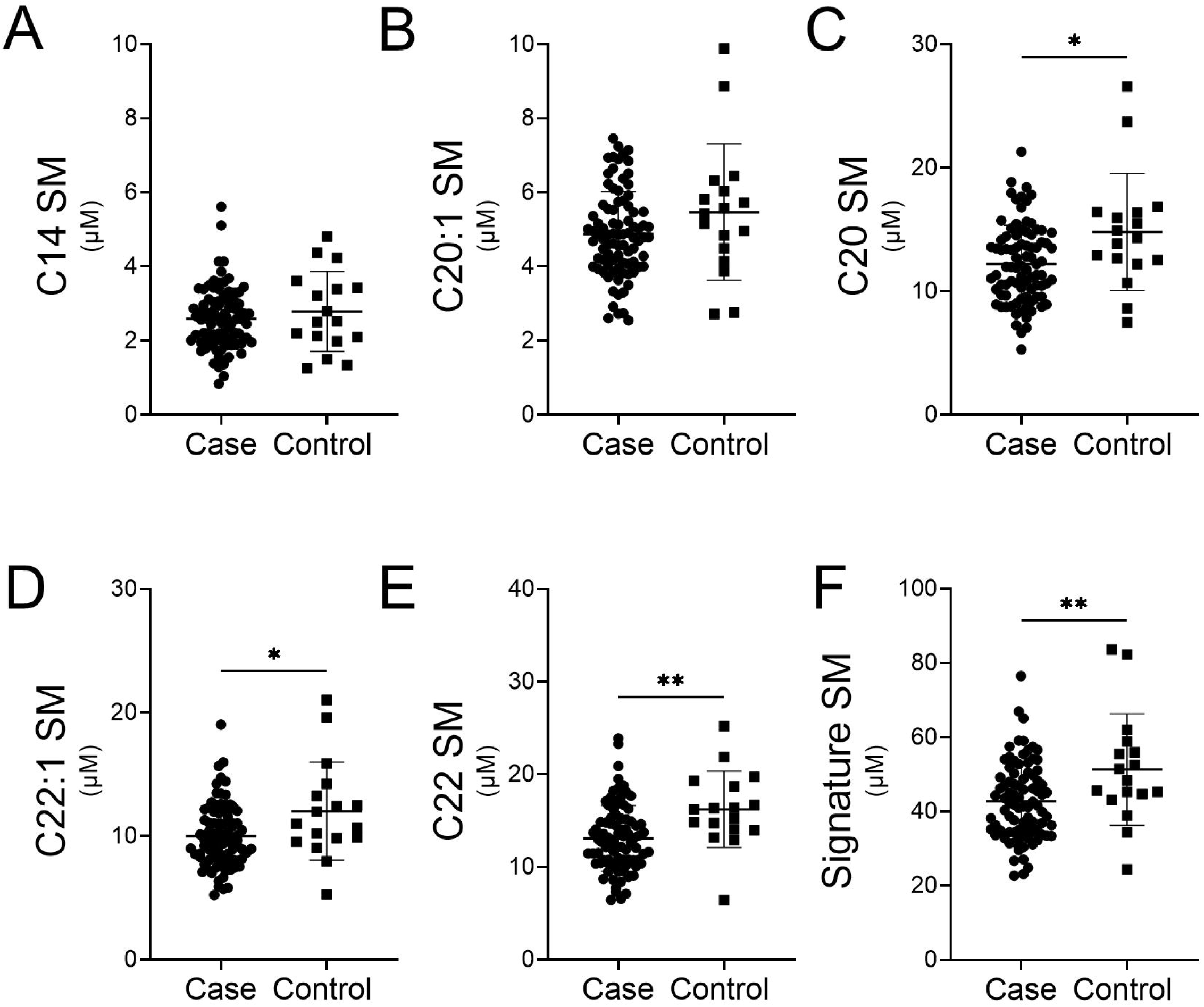
Signature SMs are decreased in CRC patients from UACC series. Lipids in serum from UACC CRC patients and clean-colon controls were extracted and analyzed for sphingolipids using LC-MS/MS. Five SM species identified in the untargeted lipidomics were quantified **A)** C14, **B)** C20:1, **C)** C20, **D)** C22:1, **E)** C22, **F)** sum of signature SMs (C14, C20:1, C20, C22:1). Data are normalized to volume and represent mean ± SD; *p<0.05, **p<0.01.

### S-SMase activity is increased in CRC cases

Decreased SM can be due to decreased generation by SM synthases (SMSs) or by increased breakdown by sphingomyelinases (SMases). Acid-SMase is found primarily in the lysosomes of cells and is also secreted extracellularly via alternative trafficking [29, 30]. The degradation of SM in serum is typically regulated by this secreted form of a-SMase, namely S-SMase [31, 32]. Therefore, we assayed S-SMase activity in serum from CRC cases from the UACC CRC case-control series to determine if increased activity correlated with decreased serum SM (samples from the CCCC CRC series had been depleted). S-SMase activity levels were significantly higher in UACC CRC patients (0.131 ± 0.098) compared to controls (0.051 ± 0.036; p <0.0001) (**Figure 3A**). Correlation analyses determined that two of the signature SMs were slightly, yet significantly, negatively correlated with S-SMase activity, namely, C22:1 SM (95% CI: -0.391 to -0.018, p=0.0323) and C22 SM (95% CI: -0.425 to -0.064, p=0.01) (**Figure S4A-F**). Multivariate analyses revealed no significant impact of age, race, or sex on S-SMase activity in the UACC CRC series (**Table S5**). These data suggested that serum S-SMase activity is increased in patients with CRC and may contribute to lower levels of serum SMs.

**Figure 3.**
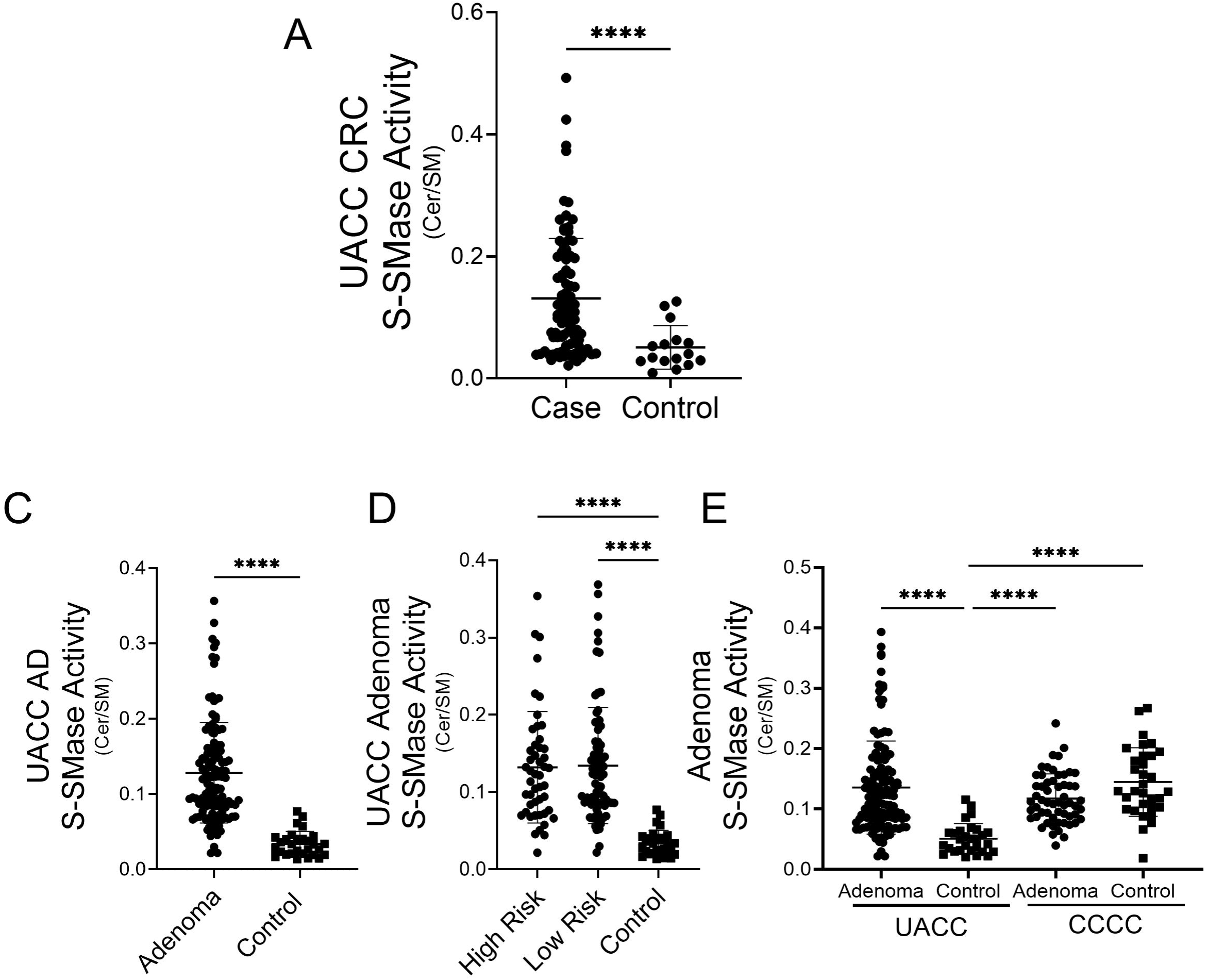
S-SMase activity is elevated in patients with adenoma and CRC. Serum S-SMase activity was analyzed from UACC adenoma, CRC patients and clean-colon controls. **A)** S-SMase activity from CRC patients and controls from the UACC series. S-SMase activity from UACC adenoma patients **C)** with any adenoma vs. control and **D)** separated based on high-risk adenoma vs. a low-risk adenoma; and **E)** S-SMase activity from UACC and CCCC control and adenoma patients. Data represent mean ± SD; ****p<0.0001.

### S-SMase activity is increased in whites with adenomatous polyps

Higher levels of S-SMase activity in serum from CRC patients compared to controls could occur in the pre-malignant phase of tumorigenesis or during cancer progression. To evaluate the potential for increased S-SMase to precede CRC development, we measured S-SMase levels in cases with non-advanced and advanced adenomas (UACC adenoma, **Table 3**). S-SMase activity levels were significantly higher adenoma cases (average ceramide/SM ratio 0.128 ± 0.067) compared to clean-colon controls (average ceramide/SM ratio 0.052 ± 0.025; p <0.0001) (**Figure 3D**). Levels of S-SMase activity were significantly increased in both case groups compared to controls but were not significantly different from each other (**Figure 3E**). Additional multivariate analyses demonstrated no significant impact of age, race, or sex on S-SMase activity in the UACC adenoma series (**Table S6**). Interestingly, S-SMase activity was not significantly different between Chicago Black adenoma cases and clean-colon controls (**Figure 3E; Table S7**). We note that the S-SMase activity in Chicago Black controls was significantly higher than in predominantly white UACC controls.

In order to further determine if the signature SMs or S-SMase activity could be used to indicate the presence or absence of adenoma or CRC, we utilized ROC curves. Analysis of individual SMs comprising the signature SMs or the signature SMs all yielded ROCs with an AUC ≤ 0.736 (**Figure S5**). ROC analysis of UACC CRC S-SMase activity resulted in a slightly higher AUC = 0.77 (**Figure 4A**); however, ROC analysis of UACC adenoma S-SMase activity resulted in an AUC =0.917 (**Figure 4B**). Together, these data suggest S-SMase activity may serve as an early detection biomarker for colorectal neoplasia.

**Figure 4.**
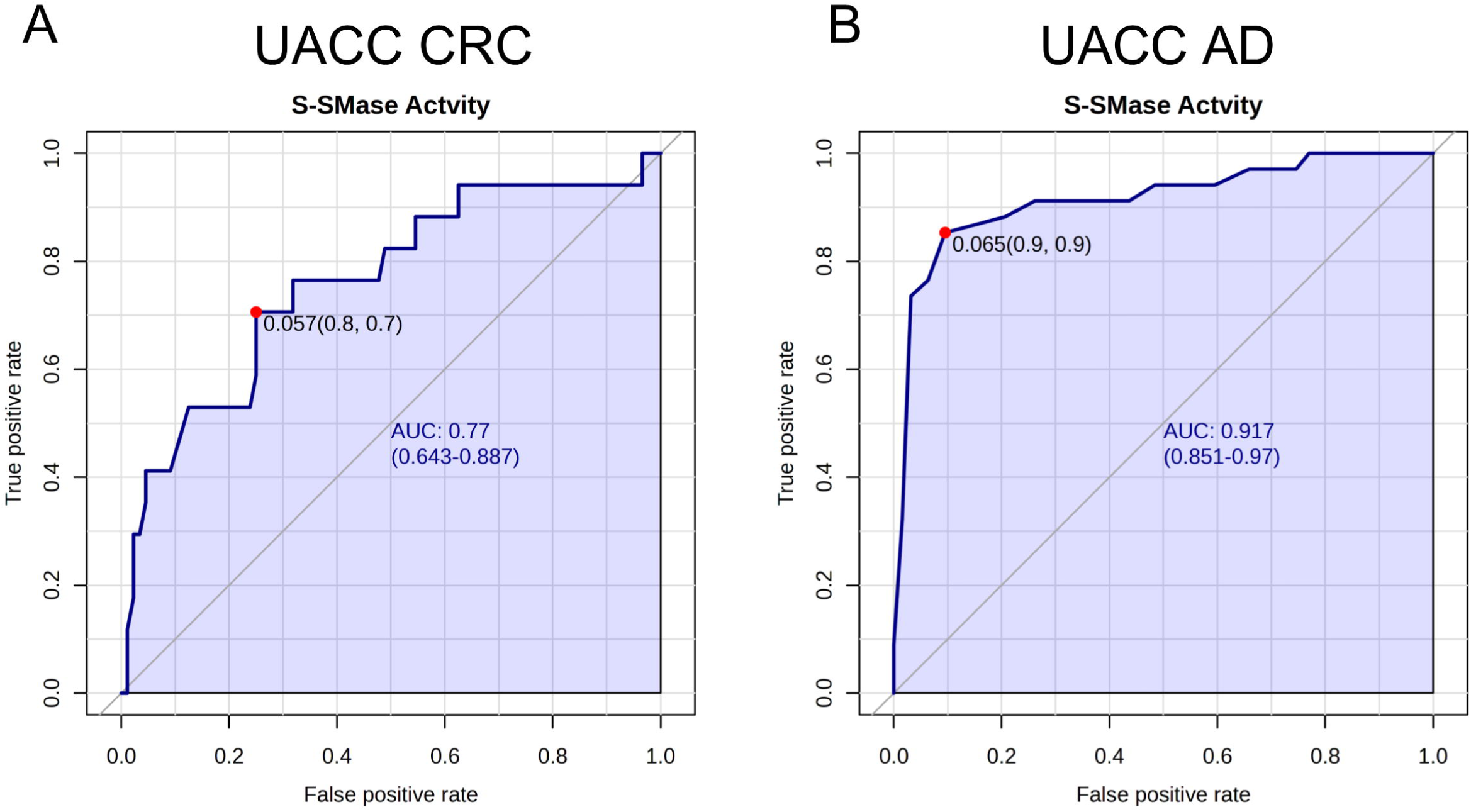
ROC analysis for S-SMase association with CRC and adenoma (AD). S-SMase activity in UACC **A)** CRC and **B)** AD were analyzed by ROC. Optimal cutoffs are indicated by the red dots.

## Discussion

In the present study, we used untargeted lipidomics in a racially diverse series of human subjects from Chicago to identify serum metabolites that were significantly different in abundance in CRC cases in comparison to clean-colon controls. Thirteen of the 48 metabolites that were identified were long-chain and very long-chain sphingolipids. Quantitative analysis using targeted LC-MS/MS showed that reduced levels of SMs were manifest in CRC cases regardless of age, sex, or race. We then substantiated these data using targeted MS analyses and S-SMase activity assays in an independent series of predominantly white CRC cases from Arizona. S-SMase levels were also elevated in patients with adenomas. As these elevated S-SMase levels were detected in patients with non-advanced adenomas, these data suggested that increased levels of S-SMase in serum may be an early event in colorectal tumorigenesis. Finally, the difference in S-SMase activity in adenoma cases compared to controls suggested that S-SMase has potential as a biomarker for detecting the presence of colorectal neoplasia, although differences were detected in white but not in black Americans.

Several studies have utilized untargeted metabolomics to generate “lipid signatures” that include species of SM to distinguish CRC samples from healthy controls. Plasma lipidomics identified seven lipid species that could be biomarkers for CRC [12], including decreased levels of two species of SM (42:2 and 38:8) in CRC samples. Untargeted lipidomics was also used to define 48 metabolites that could distinguish CRC patients from controls, one of which was C22:3 SM [33]. Several species of SM were also decreased between control patients and advanced adenomas, as well as between control and CRC patients with C22:1 SM being the only similar species between our studies [15]. Targeted analyses of sphingolipids in a small series of stage IV CRC patients (n=10 each case vs. control) also demonstrated decreased levels of C24 SM in serum [34]. These studies support our data demonstrating decreased SMs in CRC patients. However, our study is the first to report (i) quantitative levels of multiple SMs in serum, (ii) correlations between elevated levels of SM in CRC cases and elevated levels of S-SMase, and (iii) elevated levels of S-SMase in serum in patients with CRC and with adenomatous polyps.

Secretion of SMase is associated with inflammatory processes and diseases that have an inflammatory component, including metabolic diseases, hepatitis C, heart disease, Alzheimer’s, inflammatory renal disease, systemic vasculitis, and others [35]. In addition, S-SMase activity has been shown to increase with age [36]; however, our studies determined no correlation between age and S-SMase activity (**Tables S5-7**). Given the range of different clinical conditions in which elevated S-SMase levels have been reported, it will be important to screen a large number of asymptomatic adults, who are colorectal cancer screening-age and younger, to determine the range of serum S-SMases levels in persons with no colorectal neoplasia.

It is possible that colorectal tumors secrete a factor(s) that regulates secretion of S-SMase by a distant organ or that cells in the tumor are responsible for the elevated serum S-SMase. A study in samples from 3 independent cohorts of CRC patients found SM levels were elevated in CRC samples compared to normal colon tissues [11]. Additionally, total SMs were increased in 3D organoid cultures as compared to 2D organoid cultures using CRC cell lines [37]. Exosomes isolated from CRC cells and plasma from CRC patients also exhibited increased levels of d18:1/16:0 SM [38]. Because our data demonstrated increased S-SMase in serum, future studies are needed to explain these seemingly disparate results.

The SMase assay is relatively simple, and could be incorporated into protocols for early detection of colorectal neoplasia in younger age persons. With additional predictive biomarkers, one can imagine S-SMase activity as a screening approach for CRC in persons under age 45, where a blood test would be part of annual check-ups, and persons with S-SMase activity over a specific threshold then being referred for colonoscopy. It could also be informative to examine S-SMase in serum of patients participating in the PLCO (prostate, lung, colon and ovarian cancer) cohort or similar cohort studies to assess the efficiency S-SMase levels in disease prediction. As S-SMase activity was not associated with adenoma in the CCCC series (**Figure 4E**), additional prospective studies may also lend insight into the impact of age, race, and sex on S-SMase activity levels and eliminate potential confounding factors associated with collection. Indeed, multivariate analyses in signature SMs identified a significant impact of collection site on SM levels (*P*<0.0001, 95% CI = [10.55, 20.55]). As noted above, S-SMase activity from UACC and CCCC adenoma controls were significantly different (p<0.0001), suggesting substantial variation in baseline S-SMase activity among racial groups. Higher levels of SMs in American Black controls compared to white controls have been reported in multiple studies, although none of these studies assessed S-SMase enzyme activity [39-41]. Our studies also bring to light the potential for factors other than S-SMase activity to regulate serum SM levels in American Blacks. Additional studies examining the impact of race on baseline and disease state lipids, may lend insight into the impacts of race and ethnicity on sphingolipid metabolism and enzyme activity.

The limitations of this study included smaller number of samples from controls for the UACC CRC series, less epidemiologic and demographic data from the Arizona hospital-based patients and insufficient patient numbers to assess SM levels and S-SMase activity as a risk factor for EO-CRC. Our studies were also not powered to detect differences between left and right-sided disease or based on stage, though the latter point may be less important based on increased S-SMase activity in the UACC adenoma series. In addition, we did not have access to data on clinical factors that may associate with SM levels, such as medications, weight, cholesterol, triglycerides, and comorbidities. Future studies will be geared at collection of additional epidemiologic and demographic data and assessment of patients at risk for EO-CRC. Additionally, though somewhat considered a strength, samples for this study were collected at multiple sites without uniform collection methods and stored frozen for different periods of time. It will be important to follow this promising initial study with a well-designed prospective study where these variables are well controlled.

In summary, we have shown that specific species of SM are decreased in CRC patients. Patients with CRC also demonstrated higher S-SMase activity than controls. This is the first study to correlate S-SMase activity with serum SM levels in CRC patients. Higher S-SMase activity was associated with the presence of adenomas in comparison to controls in a predominantly white population. Future studies will be geared towards the development of S-SMase activity and serum SM levels as risk assessment tools for early detection of colorectal neoplasia.

## List of Abbreviations

CRC: colorectal cancer
CCCC: Chicago Colorectal Cancer Consortium
UACC: University of Arizona Cancer Center
aSMase: acid sphingomyelinase
S-SMase: secretory acid sphingomyelinase
SM: sphingomyelin
EO: early onset
USPSTF: United States Preventative Services Task Force
TARGHETS: Tissue Acquisition Repository for Gastrointestinal and Hepatic Systems
IS: internal standard
QC: quality control
UPLC: Ultra-high performance liquid chromatography
ESI: Electron spray ionization
Q-TOF-MS: Quadrupole Time-of-Flight Mass Spectrometry
LC-MS/MS: liquid chromatography–tandem mass spectrometry
FDR: false discovery rate
TAG: triacylglycerol
DAG: diacylglycerol
PE-Cer: ceramide phosphoethanolamines
AD: adenoma
PLCO: prostate, lung, colon and ovarian cancer

## ACKNOWLEDGEMENTS

The authors would like to thank the Chicago Colorectal Cancer Consortium and the University of Arizona Cancer Center for the procurement and distribution of serum samples. We also thank the University of Colorado Proteomics and Metabolomics Facility for untargeted lipidomic analyses. In addition, we thank the University of Arizona Cancer Center Analytical Chemistry Shared Resource for targeted lipidomic and S-SMase activity analyses, Carol Kepler in Tissue Acquisition and Cell and Molecular Analysis Shared Resource for providing sera from colorectal cancer patients and participants in the selenium colorectal adenoma prevention trial, Palash Mallick in University of Arizona Health Sciences Tissue Acquisition and Repository for Gastrointestinal and Hepatic Systems for providing sera from clean-colon controls, David Harris and Lawrence Mandarino in the University of Arizona Health Sciences Repository for providing sera from emergency room controls, and the Biostatistics and Bioinformatics Shared Resource for statistical expertise. The authors thank the staff, advisory committees, and research subjects participating in these studies for their important contributions.

## AUTHORS’ CONTRIBUTIONS

JMS: conceptual and experimental design, generation of MS data, analysis for experiments and wrote the manuscript. EKB, MLH, and AH: generation S-SMase activity data. QW: analysis of data. JLM, XL, RMX and PL: provided expertise and resources. ETJ: provided statistical expertise and resources. JMS, PL, NAE and AJS: conceived the original hypothesis, provided necessary funding for the materials and methods, and supervised the project. All authors contributed to editing of the manuscript.

## ETHICS APPROVAL

These studies were conducted according to the corresponding approved IRB protocols at each institution. Serum from Chicago Colorectal Cancer Consortium, colorectal and and adenoma series, were obtained previously under the protocol approved by the University of Illinois – Chicago IRB (Protocol Number 2010-0168). Serum from the University of Arizona Cancer Center colorectal series were obtained previously under protocols approved by the University of Arizona IRB (Protocol Numbers 1909985869, 0600000609, 1410545697). Serum samples from the University of Arizona Cancer Center adenoma series were obtained previously under the National Institute of Health Clinical Trials.gov number NCT00078897.

## CONSENT FOR PUBLICATION

Not applicable.

## DATA AVAILABILITY

All data are available from the corresponding authors on request.

## COMPETEING INTERESTS

The data described in the manuscript are being included in a provisional patent application.

