## Supplemental Tables for "Secretory acid sphingomyelinase activity is elevated in persons with colorectal neoplasia"

**Table S1. Top Metabolites altered in CCCC CRC series.**

| Metabolite | p-value | | Metabolite | p-value |
|---|---|---|---|---|
| 14Z,17Z,20Z,23Z,26Z,29Z-dotriacontahexaenoate | 7.75E-10 | | **Sphingomyelin (d18:1/20:0)** | 0.000114 |
| Pitheduloside I_2 | 3.78E-09 | | Bovinic acid | 0.000135 |
| Hyperforin | 1.56E-08 | | TG (14:0/14:1_9Z/18:3_9Z,12Z,15Z) | 0.000192 |
| Barringtogenol C | 2.40E-08 | | DG (15:0/18:3_9Z,12Z,15Z/0:0)_2 | 0.000298 |
| Pitheduloside I_1 | 1.53E-07 | | **Sphingomyelin (d18:1/20:1)** | 0.000338 |
| octaethyleneglycol monododecyl ether | 1.15E-06 | | DG (15:0/18:3_9Z,12Z,15Z/0:0)_3 | 0.000349 |
| ubiquinol-7 | 1.38E-06 | | Biliverdin | 0.00036 |
| PSO (20:0/22:6)_2 | 3.32E-06 | | **Sphingomyelin (d18:1/18:2)** | 0.000485 |
| O-Phosphothreonine | 5.57E-06 | | **Sphingomyelin (d18:1/14:0)** | 0.000528 |
| **FMC-5 (d18:1/20:0)** | 6.67E-06 | | Mulberrofuran E | 0.000538 |
| TG (14:0/14:1_9Z/20:3n6)_2 | 6.87E-06 | | **Sphingomyelin (d18:1/23:0)** | 0.000608 |
| DG (15:0/18:3_9Z,12Z,15Z/0:0)_1 | 8.68E-06 | | Eplerenone_2 | 0.000864 |
| **Sphingomyelin (d18:1/22:1)** | 1.08E-05 | | **Ceramide (d18:1/23:1)** | 0.001127 |
| **PE-Ceramide (d14:2/21:0)** | 1.43E-05 | | PC-P (20:0/22:6_4Z,7Z,10Z,13Z,16Z,19Z) | 0.001162 |
| 9,10-epoxyoctadecanoic acid_1 | 1.46E-05 | | 3-demethylubiquinol-8 | 0.001254 |
| **PE-Ceramide (d18:0/15:1)** | 1.84E-05 | | 3-methoxy-4-hydroxy-5-all-trans-heptaprenylbenzoic acid | 0.001612 |
| PS-P (16:0/19:0) | 1.92E-05 | | 17-4-hydroxyphenylheptadecanoate | 0.001757 |
| PS (20:0/18:0)_1 | 4.07E-05 | | 4-amino-5-hydroxy-3-all-trans-hexaprenylbenzoate | 0.002105 |
| PS-P (16:0/18:0) | 4.43E-05 | | **Ceramide-1-phospate (d18:1/14:0)** | 0.002198 |
| Dihyroxy-1H-indole glucuronide I | 5.14E-05 | | 4b-Hydroxycholesterol | 0.002204 |
| **FMC-5 (d18:1/18:0)** | 5.74E-05 | | PA-O (16:0/20:1_11Z) | 0.002231 |
| PA-O (16:0/20:2_11Z,14Z) | 5.78E-05 | | **Sphingomyelin (d18:1/22:0)** | 0.002314 |
| isovaleryl diethylamide | 7.07E-05 | | PE-P (18:0/20:1_11Z) | 0.00254 |
| trans,octacis-decaprenylphosphoryl-D-2'-keto-erythro-pentofuranose | 8.19E-05 | | Loroxanthin ester/ Loroxanthin dodecenoate | 0.003108 |

| Variable | Estimate | Standard Error | 95% CI | P-value |
|---|---|---|---|---|
| **Age** | -0.201 | 0.102 | -0.403 to 0.001 | 0.052 |
| **Sex [Female]** | 6.285 | 2.308 | 1.707 to 10.86 | *0.008 |
| **Race [B]** | -3.232 | 2.137 | -7.473 to 1.008 | 0.134 |
| **Case-Control [Case]** | -5.807 | 2.162 | -10.10 to -1.518 | *0.009 |

**Table S2. Multivariate analyses of Signature SMs in serum from CCCC CRC series.**

**Table S3. Sphingolipid levels in sera from UACC CRC series.**

| SPHINGOLIPID | CASE | CONTROL | P-VALUE |
|---|---|---|---|
| ***Sphingoid Bases*** | | | |
| Sphingosine | 0.255 ± 0.099 | 0.218 ± 0.040 | 0.324 |
| Sphinganine | 0.065 ± 0.059 | 0.047 ± 0.017 | 0.183 |
| Sphingosine-1P | 2.382 ± 1.279 | 1.315 ± 0.616 | *<0.0001 |
| Sphinganine-1P | 0.235 ± 0.154 | 0.128 ± 0.048 | *<0.0001 |
| ***Ceramides*** | | | |
| C14 Ceramide | 0.194 ± 0.062 | 0.199 ± 0.055 | 0.780 |
| C16 Ceramide | 3.689 ± 0.912 | 3.487 ±1.197 | 0.432 |
| C18:1 Ceramide | 0.601 ± 0.087 | 0.298 ± 0.077 | 0.675 |
| C18 Ceramide | 0.913 ± 0.385 | 0.721 ± 0.287 | 0.087 |
| C20:1 Ceramide | 0.045 ± 0.011 | 0.043 ± 0.013 | 0.315 |
| C20 Ceramide | 0.732 ± 0.190 | 0.658 ±0.151 | 0.207 |
| C22:1 Ceramide | 0.526 ± .0161 | 0.445 ±0.090 | *0.036 |
| C22 Ceramide | 5.505 ± 1.780 | 5.240 ± 0.949 | 0.911 |
| C24:1 Ceramide | 9.352 ± 2.573 | 8.532 ± 1.589 | 0.368 |
| C24 Ceramide | 18.61 ± 6.067 | 18.10 ± 4.090 | 0.738 |
| ***Sphingomyelins*** | | | |
| C16 Sphingomyelin | 33.71 ± 6.50 | 35.31 ±9.80 | 0.390 |
| C18:1 Sphingomyelin | 9.001 ± 2.659 | 9.619 ± 3.380 | 0.471 |
| C18 Sphingomyelin | 20.49 ±6.305 | 21.65 ± 6.648 | 0.362 |
| C24:1 Sphingomyelin | 22.85 ± 5.869 | 27.26 ± 6.397 | *0.003 |
| C24 Sphingomyelin | 8.065 ± 1.982 | 9.824 ±2.338 | *0.0009 |

| Variable | Estimate | Standard error | 95% CI | P-value |
|---|---|---|---|---|
| **Age** | -0.022 | 0.089 | -0.199 to 0.155 | 0.810 |
| **Sex [Female]** | 8.556 | 2.346 | 3.897 to 13.21 | *<0.0001 |
| **Race [AI]** | 1.113 | 9.113 | -16.98 to 19.21 | 0.903 |
| **Race [M]** | -5.745 | 6.454 | -18.56 to 7.068 | 0.376 |
| **Race [B]** | 13.830 | 8.956 | -3.957 to 31.61 | 0.126 |
| **Race [H]** | 0.022 | 3.747 | -7.417 to 7.461 | 0.995 |
| **Affection status [Case]** | -7.178 | 4.537 | -16.19 to 1.831 | 0.117 |

**Table S4. Multivariate analyses of signature SMs in serum from UACC CRC series.**

| Variable | Estimate | Standard error | 95% CI | P-value |
|---|---|---|---|---|
| **Age** | 0.000 | 0.001 | -0.001 to 0.002 | 0.538 |
| **Sex [Female]** | -0.006 | 0.016 | -0.038 to 0.025 | 0.688 |
| **Race [AI]** | 0.034 | 0.060 | -0.085 to 0.153 | 0.572 |
| **Race [M]** | 0.010 | 0.045 | -0.080 to 0.101 | 0.819 |
| **Race [B]** | -0.021 | 0.059 | -0.139 to 0.0956 | 0.716 |
| **Race [H]** | 0.004 | 0.025 | -0.045 to 0.053 | 0.861 |
| **Affection status [Case]** | 0.065 | 0.030 | 0.006 to 0.125 | *0.032 |

**Table S5. Multivariate analyses of S-Smase activity in serum from UACC CRC series.**

| Variable | Estimate | Standard error | 95% CI | P-value |
|---|---|---|---|---|
| **Age** | 0.001 | 0.001 | -0.000 to 0.002 | 0.135 |
| **Sex [Female]** | -0.005 | 0.012 | -0.029 to 0.019 | 0.680 |
| **Race [H]** | -0.005 | 0.027 | -0.059 to 0.049 | 0.849 |
| **Race [A]** | 0.021 | 0.068 | -0.114 to 0.157 | 0.755 |
| **Risk [High]** | 0.092 | 0.016 | 0.060 to 0.124 | <0.0001 |
| **Risk [Low]** | 0.092 | 0.016 | 0.060 to 0.124 | <0.0001 |

**Table S6. Multivariate analyses of S-SMase in serum from UACC adenoma series.**

| Variable | Estimate | Standard error | 95% CI | P-value |
| --- | --- | --- | --- | --- |
| **Age** | 0.000 | 0.001 | -0.002 to 0.001 | 0.767 |
| **Sex [Female]** | -0.021 | 0.015 | -0.050 to 0.008 | 0.161 |
| **Risk [High]** | -0.011 | 0.018 | -0.047 to 0.024 | 0.530 |
| **Risk [Low]** | -0.016 | 0.018 | -0.051 to 0.020 | 0.378 |

**Table S7. Multivariate analyses of S-SMase in serum from CCCC adenoma series.**
