## Supplemental Figure 1 for "Secretory acid sphingomyelinase activity is elevated in persons with colorectal neoplasia"

### Figure S1.

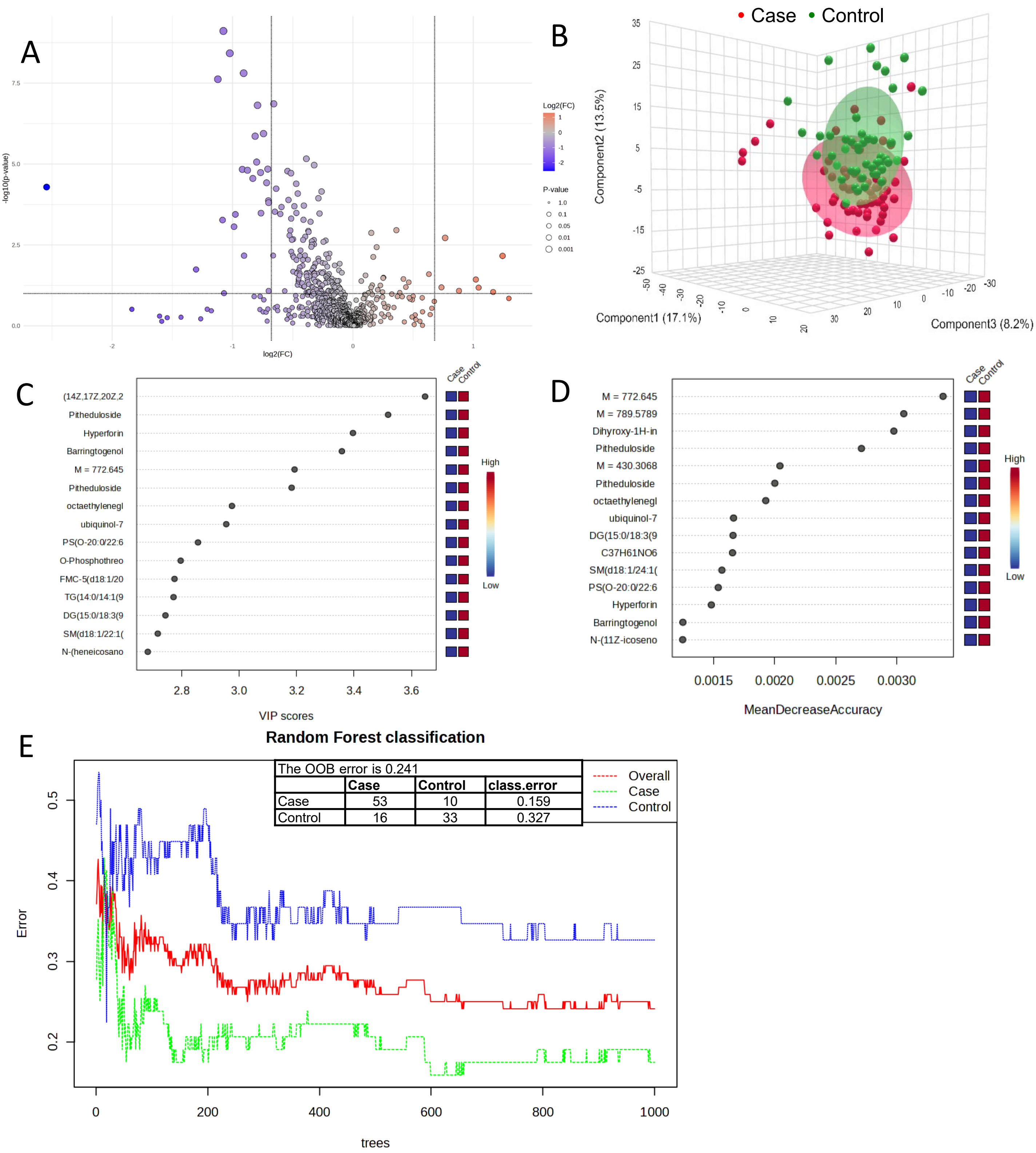


| | Case | Control | class.error |
|---|---|---|---|
| Case | 53 | 10 | 0.159 |
| Control | 16 | 33 | 0.327 |

**Figure S1. Untargeted lipidomic analyses from CCCC CRC patient serum.** Serum samples from CRC patients or clean-colon controls were subjected to untargeted lipidomic analyses. Data were analyzed using Metabolanalyst. A) Volcano plot, B) PCA, C & D) PLSDA and E) Random Forrest plots were utilized to identify lipids significantly altered in CRC patients compared to clean-colon controls.
