## Supplementary figures and images for "Secretory acid sphingomyelinase activity is elevated in persons with colorectal neoplasia"

### Supplemental Figure 2

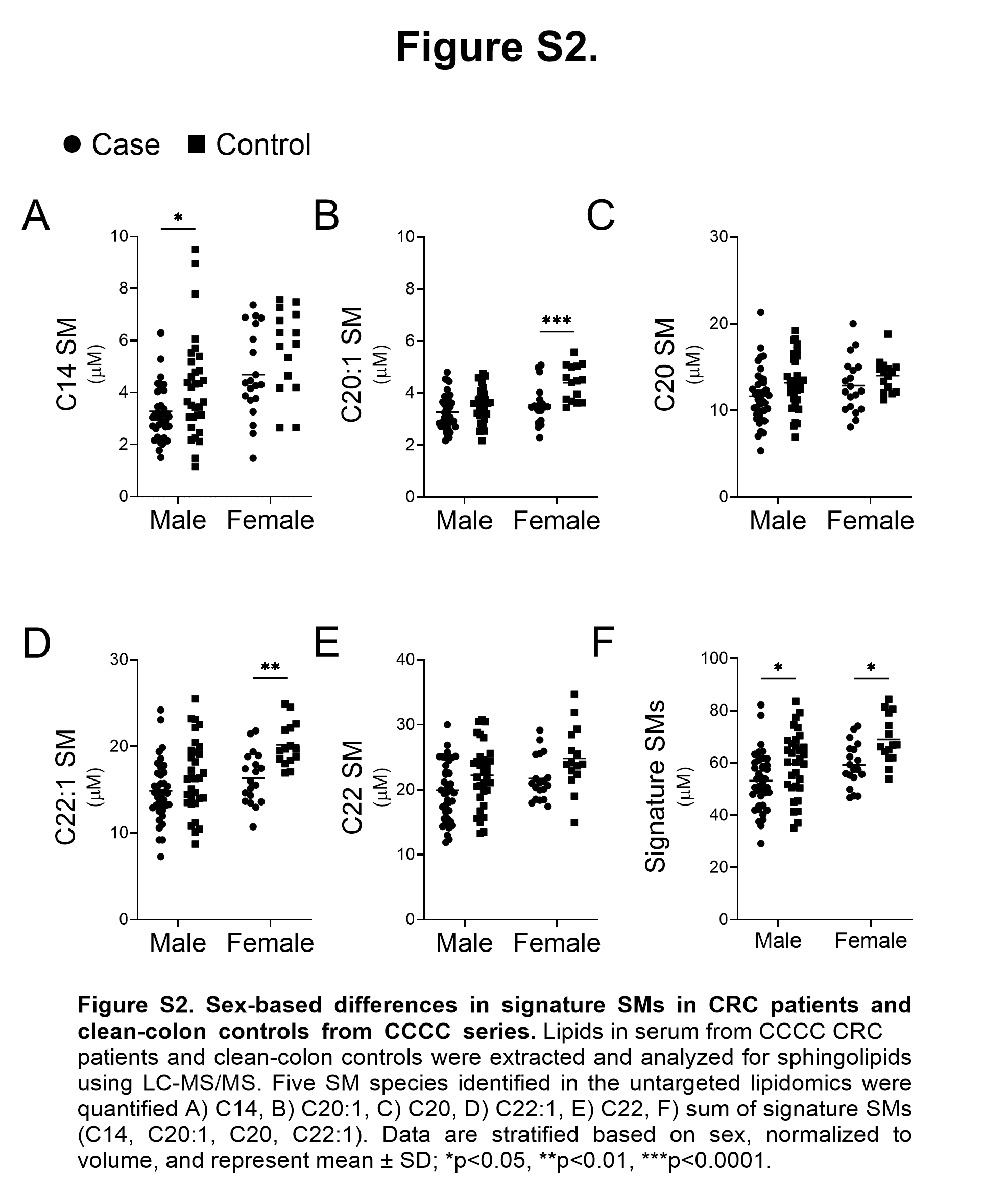

### Supplemental Figure 3

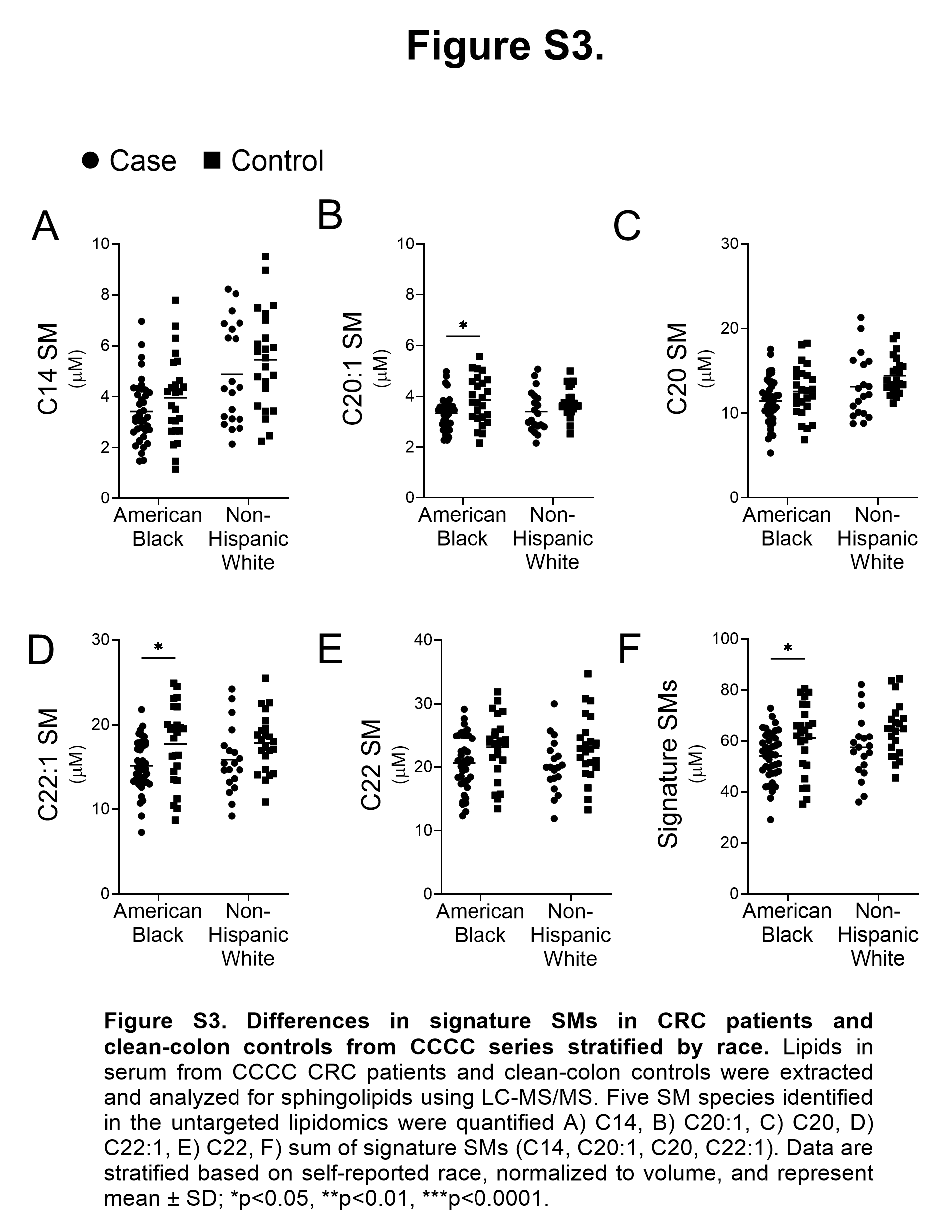

### Supplemental Figure 4

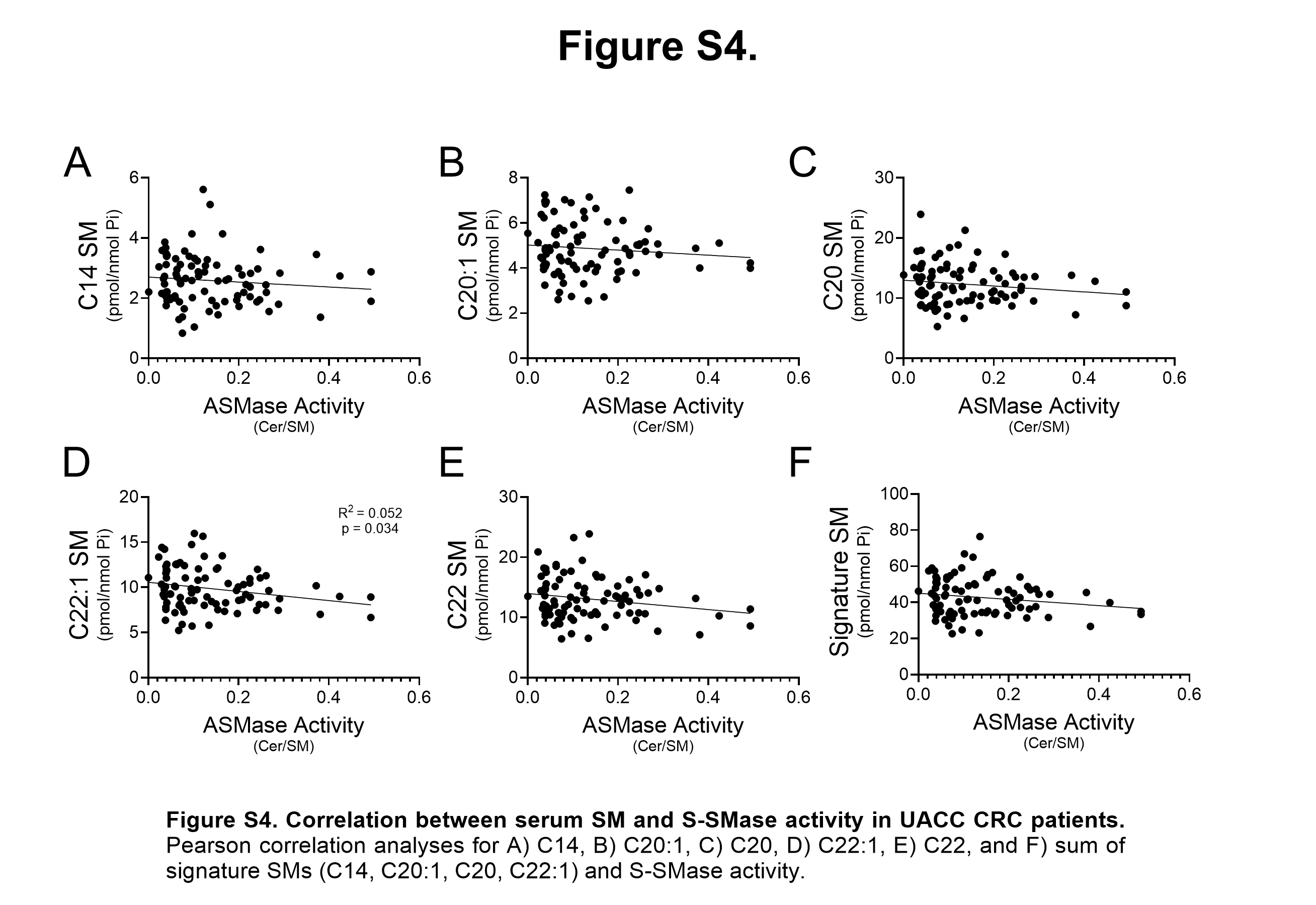

### Supplemental Figure 5

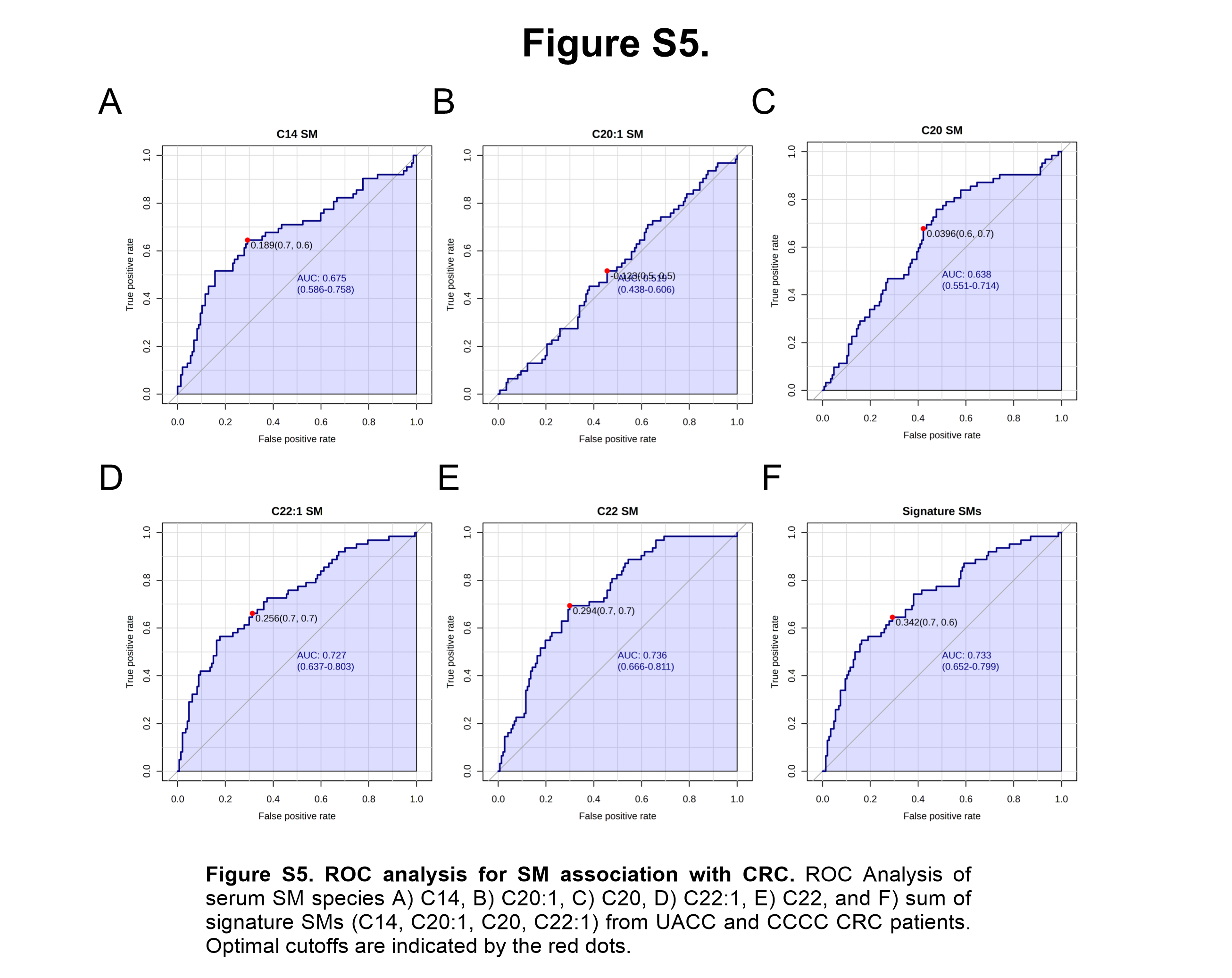
